# Wastewater surveillance to track a generational scale outbreak of measles in Ontario, Canada, February - November, 2025

**DOI:** 10.64898/2026.01.10.26343798

**Authors:** Ryland Corchis-Scott, Élisabeth Mercier, Edgard M. Mejia, Qiudi Geng, Ethan Harrop, Ana Podadera, Natalie Lewoc, Kenneth K.S. Ng, Nataliya Santiago, Natalie Knox, Lawrence Goodridge, Chand S. Mangat, Chrystal Landgraff, Karen B. Riddell, Mehdi Aloosh, Robert Delatolla, R. Michael McKay

## Abstract

The Province of Ontario (Canada) experienced a generational scale outbreak of measles in 2025. We applied wastewater surveillance concurrently with clinical-based surveillance to track measles incidence in southwestern Ontario adjacent to the United States. Measles virus (MeV) signal in wastewater was positively associated with clinical cases but did not provide early alert of changes in measles incidence when resolved by epidemiological week. Assessment of virus partitioning showed MeV RNA was broadly distributed in the liquid phase but is most concentrated in the solids. An assay was adapted for differentiation of vaccine and wildtype MeV and used to detect vaccine genotype measles following an inoculation campaign targeting underserved groups in the region. MeV shedding in wastewater was estimated through repeated sampling of sewer laterals serving a hospital treating confirmed measles infections. This measles outbreak serves as a case study highlighting the application of wastewater surveillance for measles while supporting method development in real-time.

## Introduction

Wastewater surveillance (WS) gained traction during the COVID-19 pandemic as an effective and unbiased means to track community infection without relying on clinical testing. Sampling from a wastewater treatment plant (WWTP) or locations upstream within a sewershed informed public health of hotspots of infection and tracked genetic variants associated with disease (1). Wastewater likewise yielded high-quality whole-genome SARS-CoV-2 sequence data (2), making it an important resource to track disease spread and a complement to clinical data to identify burden and potential routes of pathogen transmission (3,4). WS has extended to multiple additional targets, including respiratory diseases (5), foodborne illness (6,7), and diseases of emerging concern, such as Mpox and highly pathogenic avian influenza (8–10).

Public health officials have long warned of the increased potential for vaccine-preventable disease outbreaks (11). COVID-19 pandemic-associated disruptions and vaccine misinformation have negatively impacted immunization campaigns, including measles vaccination programs (12–14). Ontario recently experienced a generational scale outbreak of measles with >2,400 confirmed cases reported between October 2024 and October 2025 (15). Canada-wide, cases exceeded 5,375 in 2025 positioning Canada as the hotspot for measles in North America. In November 2025, following 12 months of sustained transmission, Canada lost measles elimination status (16), a designation attained in 1998 (17). The 2025 outbreak of measles in Canada likely resulted from reduced vaccination coverage (18) and was the largest Canadian outbreak in over 3 decades, overwhelming resources of public health units, especially in southwestern Ontario, which experienced the highest caseload (19).

While there was a robust public health response, measles outbreaks are costly, demanding resources and personnel to mitigate disease spread and treat patients (20). With concurrent outbreaks in the American Southwest (21) and in Alberta, Canada (22), there has been renewed interest in the application of WS to complement clinical testing (23). WS accounts for community members who may be hesitant to seek medical care (24). This is particularly relevant to measles, considering the majority of cases in North America have been linked to close-knit communities whose members are largely unvaccinated (25). Recent efforts highlight the application of WS to track measles at a community level (26–28) and the use of assays that differentiate between wild type and vaccine genotypes (23,29,30). Despite these early successes, many challenges persist to broader and more actionable adoption, including the clinical dynamics of viral shedding, viral partitioning within wastewater, and detection sensitivity (24), as well as the proximity of cases to monitored sewersheds. Here, we present a case study from a rural border municipality in southwestern Ontario demonstrating the application of WS for MeV during a measles outbreak. This event, which persisted for >9 months, supported method development and provided insights on MeV partitioning and on rates of viral shedding in wastewater.

## Methods

### Sample Collection

Composite (24-h) wastewater samples were collected 3× weekly from the Leamington Pollution Control Centre (LPCC) in the Windsor-Essex region of Ontario, Canada, between February – November 2025. The LPCC serves the urban center of Leamington and accepts sewage from residential septic tanks and storage tanks of congregate living facilities in the rural area, bringing the population served to >30,000.

Sewer laterals draining Windsor Regional Hospital (WRH) were monitored using passive samplers (tampons) deployed 1-2× weekly. Passive samplers were submerged in the hospital effluent stream for ∼17-24h. At collection, they were transported to the laboratory for immediate processing (31).

### Sample Processing for MeV Monitoring

Wastewater influent samples were concentrated by filtration or affinity capture methods, while hospital effluent samples were concentrated via centrifugation. Following concentration, total nucleic acids were extracted, and viral concentrations were measured with quantitative reverse transcription PCR (qRT-PCR). An assay targeting the MeV nucleoprotein (N) gene was used for measles quantification (Supplementary Table 1). A separate assay was used to measure pepper mild mottle virus (PMMoV) in samples (31). To validate the identity of the amplicons obtained from the N-gene of MeV, qRT-PCR products obtained from MeV-positive wastewater samples were sequenced (Supplementary Materials).

### Partitioning of endogenous MeV in wastewater

Partitioning experiments were conducted at the University of Ottawa. To determine the partitioning of endogenous MeV in wastewater, a 4 L post-grit wastewater sample collected from LPCC was processed to derive three fractions: solids, colloids, and supernatant (Figure 1). Partitioning calculations were performed using three different assumptions; a whole-sample basis, which considered wastewater composition, an equal-mass basis, and an operational volume basis, which reflects the actual processing volumes commonly used in laboratory workflows (Supplementary Materials).

**Figure 1.**
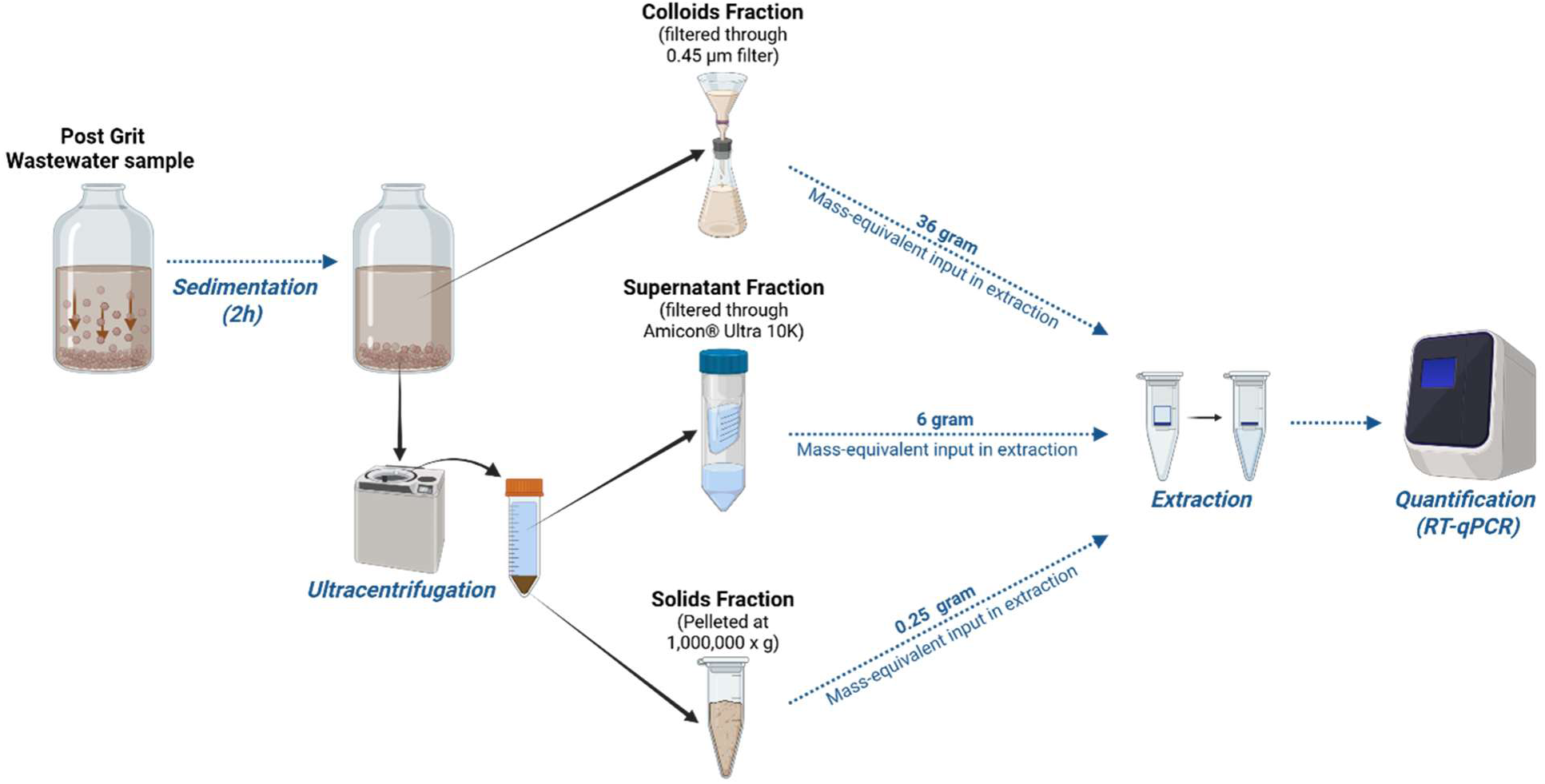
Schematic overview of the sample processing and fractionation protocol used to partition endogenous MeV into solids, colloids, and supernatant fractions in wastewater.

### Detection of vaccine genotype MeV

Aliquots of raw influent samples were shipped weekly to the National Microbiology Laboratory (NML, Winnipeg, Canada) for additional analysis. Samples were processed as previously described (32), with minor modifications. To measure MeV wild-type and vaccine genotypes in wastewater, a multiplex assay developed for clinical detection was employed. This triplex assay targets the L gene for pan-MeV detection, and the H gene for wild-type and vaccine differentiation using two locked nucleic acid (LNA) probes for respective targets (33). (Supplementary Materials).

### Additional data sources

De-identified hospital admission and discharge dates for patients admitted with measles and hospital water usage data were provided by the WRH. Weekly measles case data were obtained from the Windsor-Essex County Health Unit (WECHU). The WECHU also provided weekly vaccination data (the number of measles-mumps-rubella (MMR) vaccines administered in a targeted vaccination campaign). LPCC provided metadata (pH, temperature, flow rate) for each sample collected and the volume of septic sewage deposited at the WWTP by seven different private haulers. Research ethics approval was provided by the Windsor Regional Hospital Research Ethics Board (REB# 25-529) and the University of Windsor Research Ethics Board (REB# 25-127).

### Estimation of viral shedding

Viral shedding rates for MeV were estimated using passive samplers deployed at WRH following a previously described approach (31) with slight modifications. Shedding rate was calculated as follows:

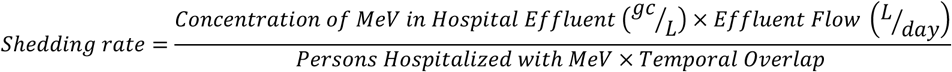

Since it is challenging to measure viral gene concentration in sampled flows using passive samplers, the concentration of MeV in hospital effluent was calculated using a previously reported mean suspended solids value for hospital effluent (34) and accounting for MeV partitioning. Flow rates were determined using hospital water use. Admission and discharge times for MeV hospitalizations were used to determine both the number of hospitalized patients and the temporal overlap value. Shedding rates are reported in log10 gene copies per day per person. The median estimated shedding rate, average weekly flow, and MeV concentration were used to create an estimated weekly case count for LPCC sewershed.

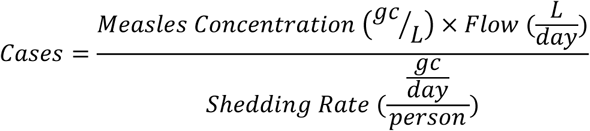

Estimated cases were compared with reported measles cases for Leamington. Additional details can be found in Supplementary Materials.

### Statistical methods

Statistical analyses were performed using R (version 4.5.1) and were restricted to data collected between February 2025 and July 2025, ending with the last reported measles case in Leamington. Spearman’s correlation was carried out between the weekly number of confirmed measles cases in Leamington and the mean weekly wastewater signal at LPCC using base R. The uncertainty of rho was estimated in the *boot* package using a non-parametric bootstrap with 1000 replications. To determine if WS for measles virus is a leading indicator, time-lagged cross correlation (TLCC) was carried out in R using the ccf boot function from the *funtimes* package. To determine which factors contributed to variability in the MeV signal in wastewater, a generalized linear model (GLM) with a gamma distribution and a log link was fitted to the data using the *glmmTMB* package in R. PMMoV normalized MeV RNA concentration was the response variable while covariates were: flow, pH, number of vaccinations administered (weekly), confirmed clinical cases, and time (epidemiological [epi]week). Predictors were standardized via mean-centering and scaled to unit variance prior to model fitting. Wastewater temperature was omitted as a covariate, since variance inflation factor (VIF) and a correlation matrix of predictors indicated that time and temperature were collinear. Model fit was checked using simulated residual diagnostics in the *DHARMa* package in R.

To investigate a potential relationship between the total volume of wastewater deposited by septic haulers and MeV wastewater concentrations, a simplified gamma GLM with a log link was fitted to a shorter data set. Normalized MeV RNA concentration was the response variable, while covariates were confirmed clinical cases and total volume of sewage deposited. Finally, correlations were run between MeV and normalized MeV concentrations, as well as the weekly septic waste volumes and daily septic waste volumes. Daily data were not included in the GLMs, as case data and vaccination administrations were resolved at weekly intervals.

## Results and Discussion

### Partitioning of MeV in wastewater

The distribution of endogenous MeV was determined across three fractions of wastewater: settled solids, colloids, and supernatant. MeV distribution was calculated using three different approaches (Figure 2). When accounting for typical wastewater composition (99.96% liquid and 0.04% solids), most of the MeV signal was detected in the supernatant (91.20 ± 7.22%), followed by the colloids (8.58 ± 1.24%), and settled solids (0.25 ± 0.05%) (Figure 2A). This approach was influenced by the disproportionate volume of the liquid phase and may not reflect the concentration potential of each matrix. On an equivalent mass basis (1g each of solids and liquids), the highest concentration of MeV was found in the solids (95.95 ± 17.75%), with lower proportions in supernatant (13.31 ± 1.05%) and colloids

**Figure 2.**
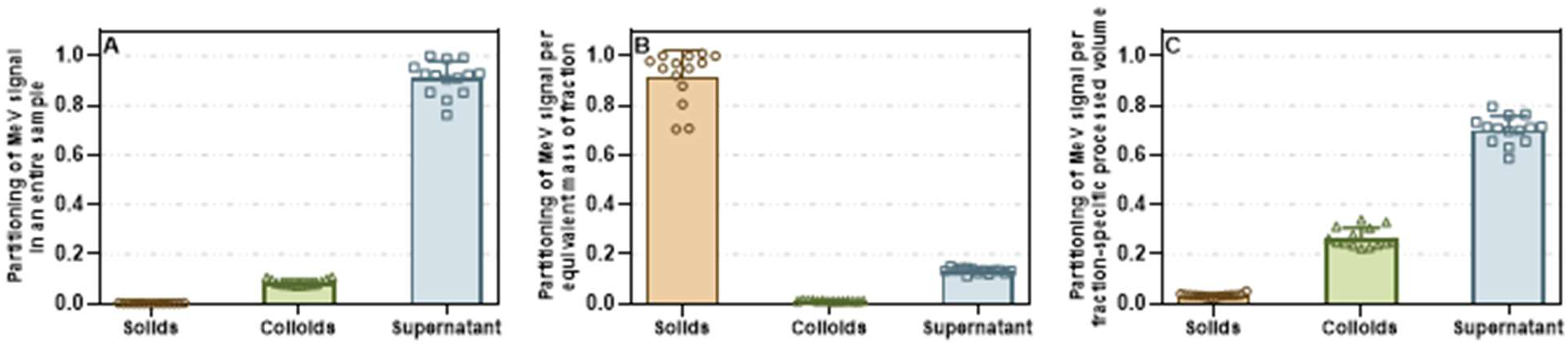
Partitioning of MeV signal across wastewater fractions under three calculation bases. (A) Signal partitioned assuming the composition of a typical wastewater sample (99.96% liquid, 0.04% solids). (B) Signal partitioned on an equivalent mass basis, assuming 1 gram of each fraction. (C) Signal partitioned based on typical laboratory processing volumes: 0.25 g for settled solids, 150 mL for the colloids fraction, and 37 mL for the supernatant. Means and standard error are displayed. Where the standard deviation is too small, the error bars are not displayed. Each point represents one technical triplicate for each of the biological replicates (n=14).

(1.25 ± 0.18%) (Figure 2B). However, routine enrichment methods do not process equivalent masses, limiting the relevance of this comparison. Weight and volume units typically processed in laboratory workflows (0.25 g for solids, 150 mL for colloids, and 37 mL for supernatant) were used for the final calculation. Under operational conditions, most of the MeV signal was recovered from the supernatant (69.92 ± 5.53%), followed by the colloids (26.67 ± 3.86%), and the solids (3.40 ± 0.63%) (Figure 2C). These findings demonstrate that MeV RNA is broadly distributed in the liquid phase but is most concentrated in the solids. However, when considering viral recovery per standard enrichment protocol, processing of the supernatant yields the highest amount of MeV RNA due to the larger volume concentrated, supporting its use for optimal detection.

### Wastewater surveillance of MeV at location of outbreak – Leamington, Ontario

WS of MeV was initiated in March 2025, following reports of cases and exposures in Windsor-Essex. Retrospective analysis of archived samples extending to February was also conducted. The first wastewater samples that tested positive for MeV were collected on March 7, 2025 (Figure 3), and samples remained conclusively positive through mid-August 2025. Normalized (ρ = 0.77, 95% CI [0.51-0.87], *p* <0.001) and raw (ρ = 0.77, 95% CI [0.49-0.90], *p* <0.001). MeV wastewater signal showed a strong positive correlation with reported cases between February 2025 and July 2025. Correlations between wastewater signal derived from a subset of samples concentrated with affinity capture techniques corroborated this result: normalized (ρ = 0.84, 95% CI [0.60-0.94], *p* <0.001) and raw (ρ = 0.87, 95% CI [0.68-0.95], *p* <0.001). The slight increase in correlation coefficient for this sample set could be a result of the truncated data set or from the use of affinity capture methods, which are more effective than filtration in concentrating MeV from the liquid fraction. The correlation exists even where populations might underutilize health services, likely contributing to the underreporting of measles cases (35). In this instance, WS for measles was robust, even though it was conducted in a rural community where a substantial portion of the population is served by septic systems. This finding suggests that WS can be an accurate, independent measure of MeV on a weekly time scale at this site. The identity of amplicons obtained by RT-qPCR was confirmed by Nanopore and Sanger sequencing. Sequence alignments using BLAST confirmed that the reads obtained shared ≥99% identity with Measles Virus nucleoprotein (N) gene.

**Figure 3.**
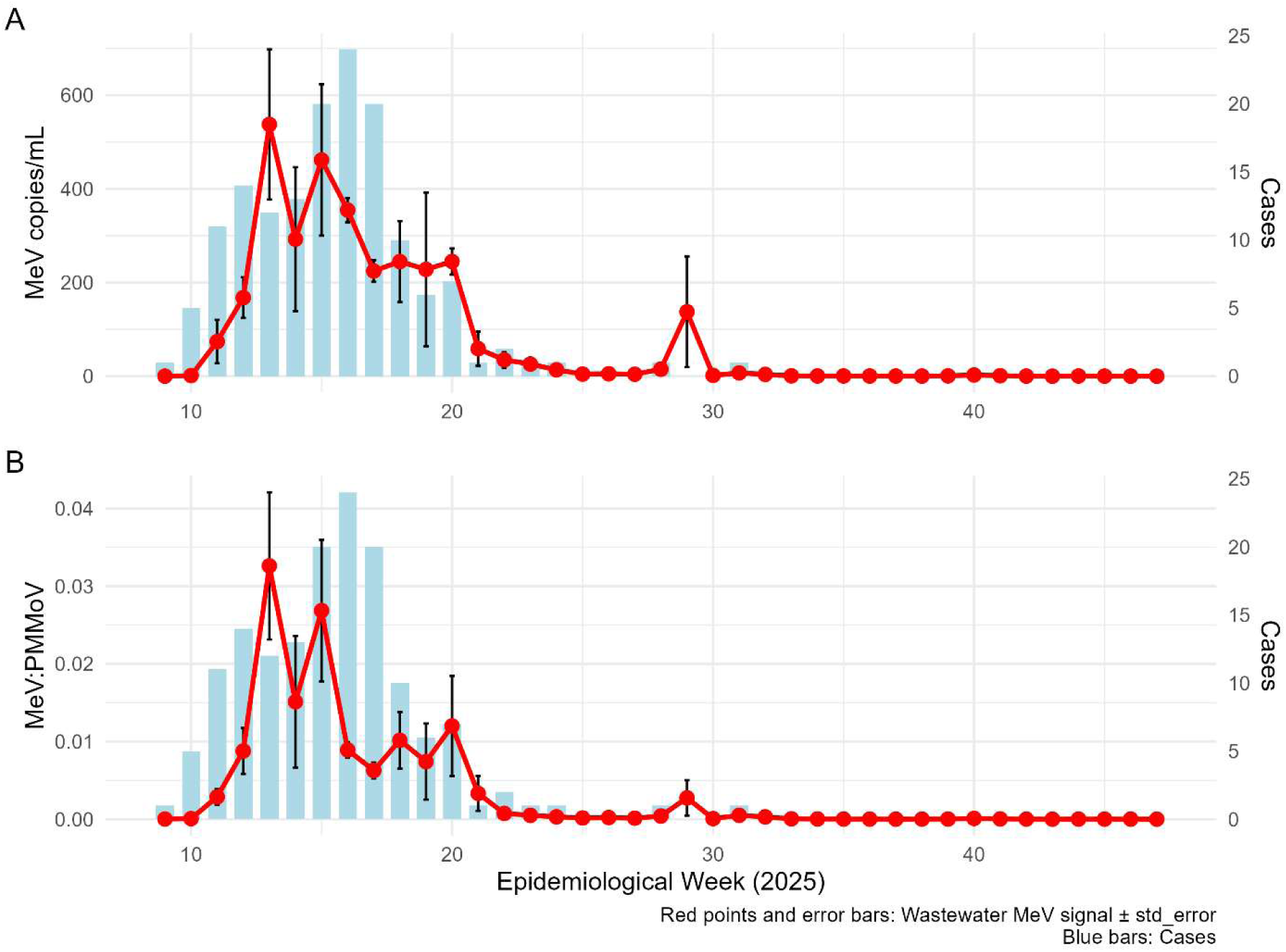
Measles virus (MeV) wastewater surveillance and clinical case counts in Leamington, Ontario (2025). Clinical cases (blue bars) are compared against **(A)** raw MeV RNA concentrations (copies/mL; red points/line) and **(B)** MeV RNA concentrations normalized to Pepper Mild Mottle Virus (PMMoV; red points/line). Data are aggregated by epidemiological week; wastewater values represent weekly means ± standard error.

WS has repeatedly been shown to be a leading indicator of disease incidence (36–40). TLCC was used to determine if WS for measles is similarly a leading indicator of clinical reporting. The maximum association was found when MeV wastewater signal lagged clinical cases by 1 week. This was consistent for both PMMoV normalized and raw signal (Supplemental Figures 1, 2). Maximum correlation values found at a lag of 1 week were not statistically significantly different from the correlations found when the time series were aligned (ρ = 0.79 and ρ =0.78 for raw and normalized signal respectively). TLCC suggests that wastewater is not a leading indicator of measles cases for data aggregated by epi-week. Weekly averaging or potential delays in the detection of MeV signals at LPCC associated with septic hauling could obscure lead-time.

WS can provide advanced warnings for changes in disease incidence during a respiratory season, prolonged outbreak or epidemic, but it can also provide warning of the onset of an outbreak or respiratory season (40, 52). During this investigation, sampling was conducted in response to reports of measles cases and thus it did not provide advanced warning of outbreak onset. Retrospective analysis of archival samples showed MeV RNA was not detected in wastewater samples prior to the first reported case. The current WS modality failed to warn of outbreak onset, potentially due to 3× weekly sampling frequency, limited method sensitivity, or the rural nature of the sewershed in which the outbreak was concentrated.

### Modelling determinants of MeV RNA concentration in wastewater samples

The gamma GLM explained 75% of the variation in normalized MeV (pseudo R^2^ = 0.75). Cases were the only statistically significant predictor. Cases had a positive influence on MeV signal (β = 2.05 ± 0.77, z = 2.67, *p =* 0.008), suggesting increased case counts correspond to greater MeV signal. An increase in cases by one standard deviation resulted in ∼8-fold increase in normalized MeV. Other fixed effects (flow, pH, epidemiological week, and vaccination rates) were not statistically significant (Table 1).

**Table 1.** Parameter estimates for the initial gamma GLM (log link) predicting normalized MeV concentration (n = 23).

| Predictor | Estimate | Std.Error | Z-value | p-value | Significance |
| --- | --- | --- | --- | --- | --- |
| Intercept | -8.28 | 2.08 | -3.99 | p < 0.001 | *** |
| Cases | 2.05 | 0.77 | 2.67 | p = 0.008 | ** |
| Flow | 0.62 | 0.41 | 1.50 | p = 0.13 |  |
| pH | 0.43 | 0.32 | 1.37 | p = 0.17 |  |
| Time | 0.12 | 0.10 | 1.17 | p = 0.24 |  |
| Vaccinations | -0.65 | 0.45 | -1.45 | p = 0.15 |  |
| Model fit: AIC = -210.3; Dispersion = 1.23; n = 23.<br>Significance codes: ***p < 0.001; **p < 0.01; p < 0.05<br>DHARMA simulated residuals showed evidence of mild heteroskedasticity (Supplementary Table 2) |  |  |  |  |  |

A separate gamma GLM was fitted to a truncated dataset (n=14), to allow the inclusion of the volume of sewage deposited by haulers at the LPCC each week. This model included cases and total sewage volume as predictors of normalized measles concentration (Table 2) and explained 72% of variability in normalized MeV concentration (pseudo R^2^ = 0.72). DHARMa simulated residuals showed evidence of mild heteroskedasticity (Supplementary Table 3).

**Table 2.** Parameter estimates for the gamma GLM (log link) predicting normalized MeV concentration fitted to the truncated data set (n = 14).

| Predictor | Estimate | Std.Error | Z-value | p-value | Significance |
| --- | --- | --- | --- | --- | --- |
| Intercept | -5.53 | 0.30 | -18.63 | p < 0.001 | *** |
| Cases | 0.79 | 0.31 | 2.58 | p = 0.01 | ** |
| Sum of septic sewage deposited | 0.90 | 0.32 | 2.86 | p = 0.004 | ** |
| Model fit: AIC = -107.6; Dispersion = 0.88; n = 14.<br>Significance codes: ***p < 0.001; **p < 0.01; p < 0.05 |  |  |  |  |  |

Analysis of the truncated dataset corroborates that cases of measles are the most significant contributing factor explaining the variability in normalized MeV concentration at the WWTP. This result reinforces the ability of WS to act as an independent measure of disease incidence within a community. The total volume of sewage deposited at the WWTP was also a significant predictor of MeV signal. Septic sewage may contain MeV RNA, contributing to the MeV signal at the WWTP. Septage deliveries would delay the arrival of viral material to the treatment facility, and thus, septic hauling likely contributes to the mild lag/loss of prediction observed for WS of measles cases at epi-week temporal granularity (Supplemental Figure 3).

Spearman correlations were run between physicochemical parameters of wastewater, MeV signal, and the total volume deposited by septic sewage hauling companies on a weekly basis. Positive correlations could be observed between the volumes deposited by select companies and MeV signal (Supplemental Figure 4). Total volume for Company 3 was particularly strongly correlated with both normalized (ρ = 0.83, *p* <0.001) and raw MeV signal in wastewater (ρ = 0.78, *p =* 0.002). A strong correlation can also be observed between the total volume of all septic sewage deposited and the raw (ρ = 0.78, *p =* 0.002) and normalized MeV signal (ρ = 0.81, *p* <0.001). These correlations indicate septic hauling may be contributing to the MeV signal at LPCC. However, the association was tested again via correlation on a dataset that only included days when septic sewage deliveries coincided with wastewater sample collection (Supplemental Figure 5). When restricted to days with coincident sampling, correlations were weaker and non-significant (Supplemental Figure 6). Thus, septic deliveries play a more minor role in explaining wastewater signal than the first correlation analysis suggested.

### Vaccine genotype detection

An assay that targets a SNP in the MeV H-gene to distinguish vaccine genotype MeV RNA from wildtype MeV RNA was also employed during this investigation (33). The detection of the SNP in the H-gene was treated as a binary indicator of the presence of vaccine genotype MeV RNA in wastewater. Vaccine genotype MeV was detected in Leamington wastewater approximately 6 weeks following the start of vaccination campaigns and approximately 4 weeks following the intensification of the vaccination campaign (Figure 4). Research in MeV shedding dynamics following vaccination indicates that shedding can start within 1 day of inoculation and can persist for up to 14 days in urine and 29 days in nasopharyngeal swabs (41,42). These values partially explain the delay between the start of the vaccination campaign and the first detections in wastewater but are unlikely to account for the full lag. In this measles outbreak, the vaccination campaign focused mainly on agri-food workers housed in bunkhouses served by septic storage tanks, and thus, the delay could be explained by the delayed delivery of MeV RNA to the WWTP. The delay indicates that the frequency of septic hauling may contribute to the temporal dynamics of WS for rural sewersheds.

**Figure 4.**
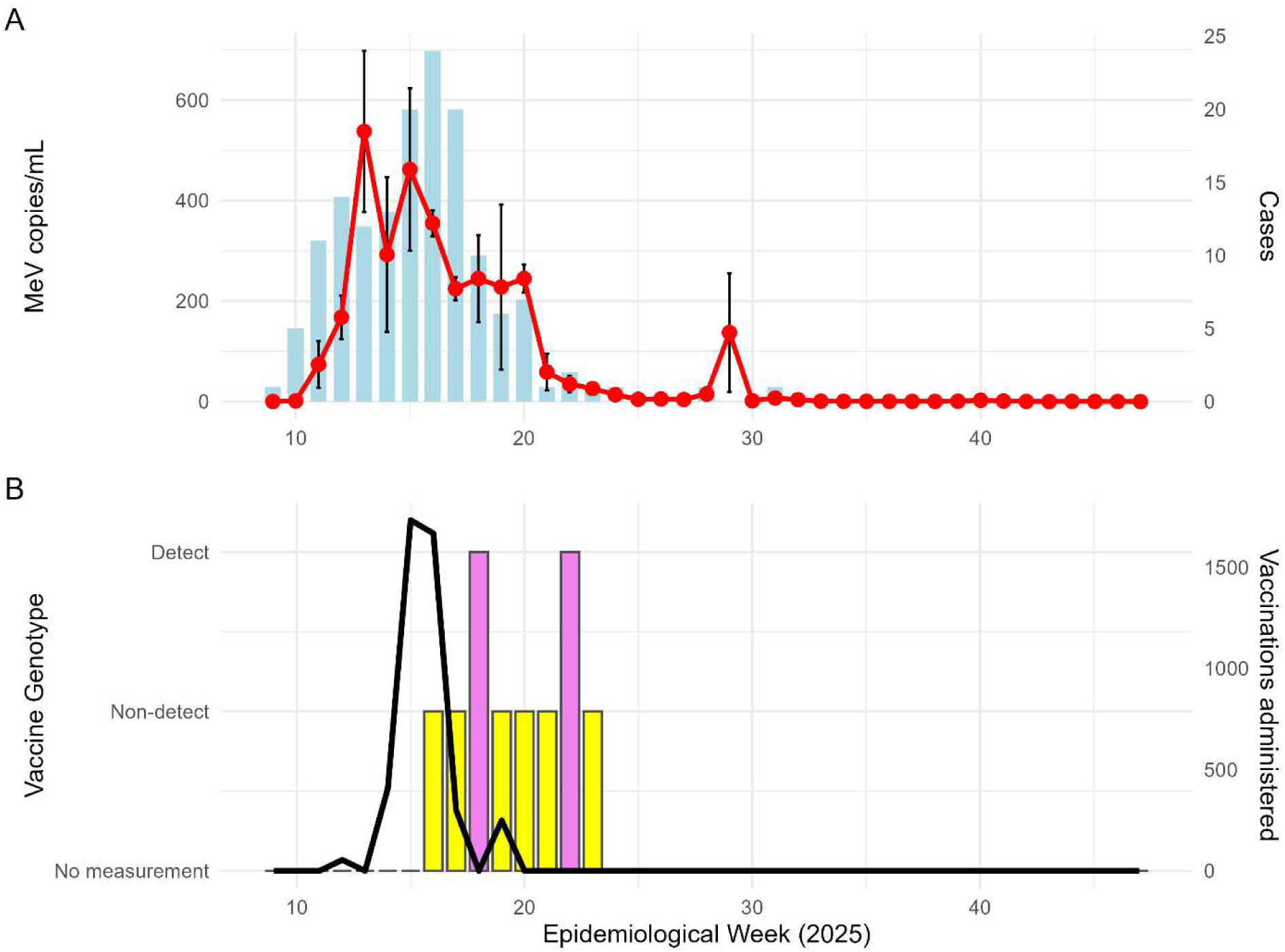
Wastewater surveillance, clinical cases, and vaccination data for measles virus (MeV) in Leamington, Ontario (2025). **(A)** Weekly clinical case counts (blue bars) compared with wastewater concentration of measles virus (copies/mL; red points). **(B)** Weekly vaccine administrations (black line) with a binary indicator (bars) of wastewater detections of MeV H-gene SNP unique to the vaccine genotype MeV. All data are displayed on an epi-week basis and represent weekly rates and mean concentrations.

### MeV Shedding and case estimation

The City of Windsor was less impacted by the measles outbreak, consistent with higher vaccination rates in urban centers (43,44). Cases and wastewater signal were both sporadic, each supporting lower incidence in Windsor (not shown). Simultaneous detections in hospital effluent at a campus of WRH and hospitalizations allowed for estimation of viral shedding rates on six dates (Table 3). Shedding rates ranged over four orders of magnitude with a median shedding rate of 11.15 log10 gene copies person^-1^ day^-1^. For one hospitalized patient, shedding was estimated on consecutive days. Shedding nominally decreased between measurements, but this decrease could be explained by measurement error. Previous studies have shown that shedding rates of measles vary significantly over the course of infection and between individuals, with a general decline in titres over time (45,46). The declining trend is not consistently supported, with another study finding no difference in MeV RNA concentration between early or intermediate oral fluid or serum samples (47). Shedding rates are variable between individuals and excreta type (48), with urine displaying relatively high MeV concentrations (24). MeV RNA levels do not differ by vaccination status or age (47,49). Our estimates of MeV shedding rates are higher than previous estimates (24). However, given that multiple sources of excreta contribute to the wastewater signal and high variability in MeV RNA measurements previously reported (45), the shedding estimates derived in this study are reasonable.

**Table 3.** MeV shedding estimates for wastewater based on samples collected from hospital effluent.

| Date | Patients Hospitalized | Shedding Estimate |
| --- | --- | --- |
| 03-25-2025 | 2 | 12.19±11.42 |
| 03-26-2025 | 2 | 12.82±11.62 |
| 04-08-2025 | 1 | 8.57±8.17 |
| 04-09-2025 | 1 | 8.52±8.20 |
| 04-22-2025 | 2 | 8.88 ±8.85 |
| 05-06-2025 | 1 | 11.62±10.99 |

Additionally, when the median of shedding rate estimates calculated here was used to extrapolate the number of measles cases in Leamington based on the wastewater signal, general agreement was observed (Figure 5). Public health officials estimate that the true incidence of measles in the Leamington outbreak was likely 5-10 times higher than reported cases (M. Aloosh, pers. comm., email, October 3, 2025). This estimate is consistent with recent MeV outbreaks in undervaccinated communities in the Netherlands (50).

**Figure 5.**
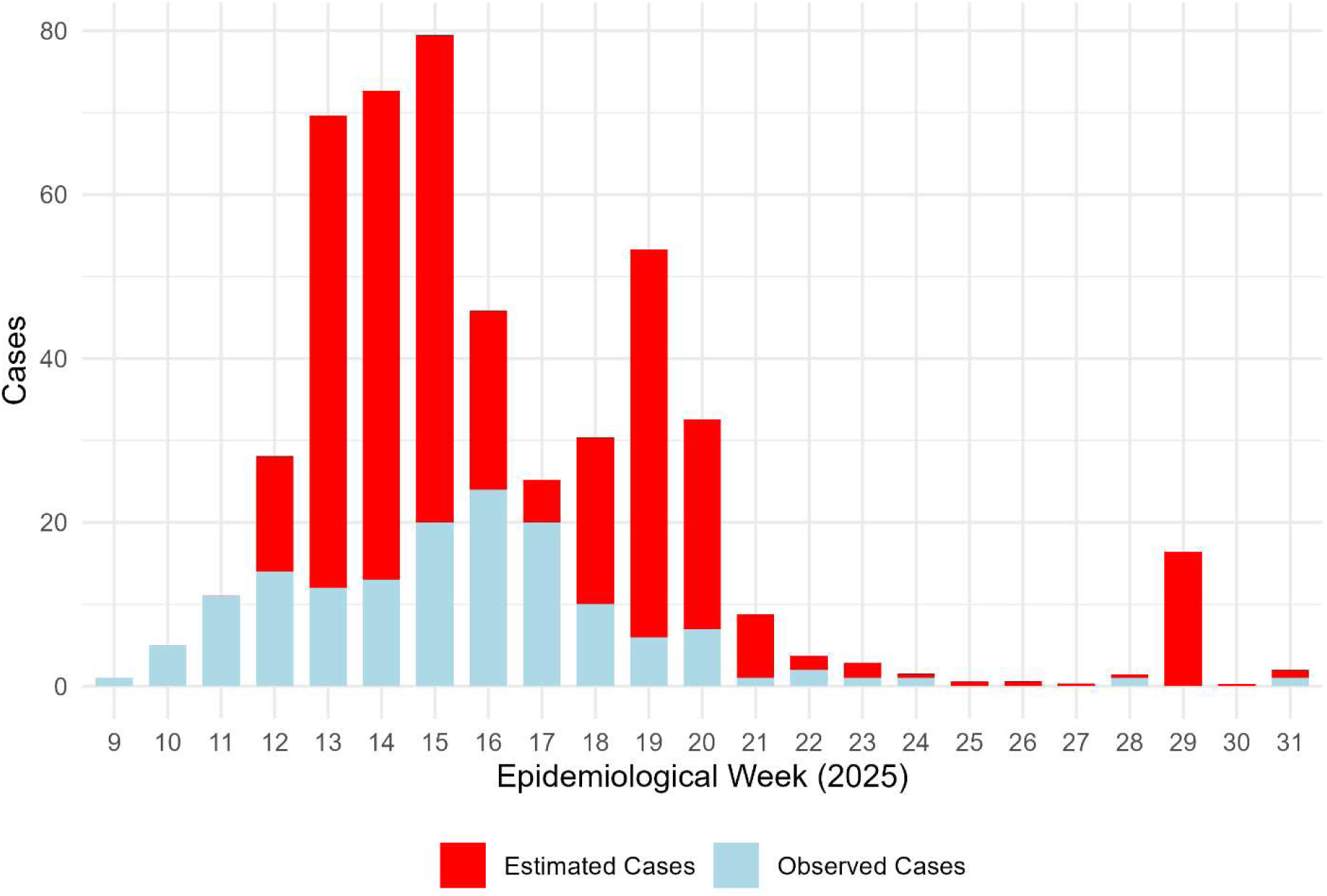
Measles case estimates for the municipality of Leamington, ON, Canada based on wastewater signal and MeV shedding approximations generated in Windsor, ON, Canada. Case estimations are based on median shedding rate approximations. Observed cases are also shown in blue, estimated cases are shown in red

## Conclusion

This investigation demonstrates that WS is an accurate independent measure of measles incidence. WS was not found to be a leading indicator of cases when data were aggregated by epi-week. However, lead-time may have been masked by weekly averaging or the influence of septic sewage haulers on delaying signal delivery to the WWTP. Septic systems and sewage hauling can influence the dynamics of measles signal in wastewater; this result could likely be extrapolated to other viruses in different rural systems with similar sewage systems. Despite the possible influence of septic hauling on the temporal dynamics of wastewater signal, WS remained an accurate and unbiased means of tracking disease at the community level. Shedding rates calculated from a sewer lateral of a hospital with confirmed measles admissions were applied to Leamington wastewater data to produce plausible estimates of measles cases.

Investigation of MeV fractionation in wastewater indicates that the majority of virus is found within the liquid phase of the wastewater matrix, consistent with measles shedding primarily in urine and previous work (30). Concentration of the supernatant may be most efficient for the purpose of WS for MeV. This investigation supports the continued use of WS for MeV, as it complements traditional surveillance for measles and remains viable in challenging circumstances where clinical metrics may under-represent actual cases. This is true despite the complications posed by septic systems.

## Limitations

Shedding rates were indirectly estimated based on mean suspended solids values for hospital effluent and MeV partitioning in wastewater and must be cautiously interpreted. Additional uncertainty in shedding rate estimates arises from flow rates being estimated using facility water usage. The LPCC serves Leamington proper and receives waste from residences served by septic systems. However, outside of the agri-food industry, most septic tanks are emptied infrequently. It is unlikely that households emptied septic tanks during the measles outbreak in a manner that contributed meaningfully to the MeV signal at the treatment facility. Also, diapering of infected children reduces the MeV load at the WWTP and potentially influences the signal.

## Data Availability

All data produced in the present study are available upon reasonable request to the authors.

## Acknowledgements

We extend appreciation to Operators and Laboratory Team with the Leamington Pollution Control Centre, Shannon Belleau, Manager of Environmental Services, Municipality of Leamington and to Kevin Douglas, Director of Facilities and Materials Management at Windsor Regional Hospital. This work was supported by INSPIRE (Integrated Network for the Surveillance of Pathogens: Increasing REsilience and capacity in Canada’s pandemic response), a program funded through the Canada Biomedical Research Fund Stage 2 (grant no. CBRF2-2023-00008), the Biosciences Research Infrastructure Fund Stage 2, and the Ontario Research Fund. Additional support was provided by the CIHR Applied Public Health Research Chair in Environment, Climate Change and One Health, awarded to Robert Delatolla.

