## Supplementary Materials for "Wastewater surveillance to track a generational scale outbreak of measles in Ontario, Canada, February - November, 2025"

### Supplementary Methods

#### Sample collection

Wastewater samples were collected 3× weekly from the Leamington Pollution Control Centre (LPCC) in the Windsor-Essex region of Ontario, Canada between February and November, 2025. The LPCC serves the urban center of Leamington and accepts sewage transported by haulers who service residential septic tanks and storage tanks of bunkhouses in the rural area, bringing the total population served to ~30,000. Wastewater samples were collected using an autosampler that composites wastewater from the influent stream at regular intervals over a 24-hour period. One litre samples were held at 4 °C for no longer than 6 days before transport in insulated coolers with ice packs to the laboratory for processing.

Upstream sampling was conducted at Windsor Regional Hospital (WRH) where sewer laterals were sampled using passive samplers deployed in the effluent stream for ~15 h at a frequency of 1-2 × weekly. Once collected, passive samplers were placed in sealable plastic bags and transported to the laboratory on ice for immediate processing. Samplers consisted of a feminine hygiene product (Tampax Cardboard Tampons, Regular Absorbency, Procter & Gamble, Cincinnati, OH, USA) attached to a carabiner which was attached to the interior of the rim of a sewer cover via fishing line and a magnet. Duplicate tampons were placed within each monitored sewer lateral to increase the volume of wastewater absorbed.

**Sample processing**

Samples of raw wastewater influent were processed at three laboratories: the University of Windsor, the University of Ottawa, and the Public Health Agency of Canada National Microbiology Laboratory (NML, Winnipeg, Canada). At Windsor, samples were concentrated by filtration using a 0.22 µm Sterivex PES cartridge filter (MilliporeSigma, Burlington, MA, USA) as described elsewhere (1). Following filtration, filter cartridges were sealed, flash frozen in liquid nitrogen and stored at -80ºC. After thawing each filter cartridge, the nucleic acids were extracted using the AllPrep Power Viral DNA/RNA kit (Qiagen, Germantown, MD, USA) modified by the addition of 5% 2-mercaptoethanol (v/v). Nucleic acids were eluted in 50 µL of RNase-free water and stored at -80°C prior to RT-qPCR and sequencing analysis. For passive samplers, collected liquids and solids were expelled manually from the feminine hygiene products and decanted into a sterile 50 mL conical polypropylene tube. The solid and liquid fractions were separated via centrifugation at 4820 × *g* for 40 min at 4°C. The supernatant was discarded, and nucleic acids were extracted from the pellet as described above.

At the NML, wastewater samples were processed as previously described (2), with a few alterations. Briefly, a pellet was recovered from a 30 mL aliquot of a 24-hour composite sample of primary post-grit influent or raw wastewater by centrifugation at 4,200 × *g* at 4°C for 22 min. Supernatant was then discarded, and the pellet was re-suspended by pipette using 700µL of buffer RLT (Qiagen) supplemented with 1% 2-mercaptoethanol (MilliporeSigma, Burlington, MA, USA). The entire volume was then transferred to a 2 mL bead-beating tube containing 200 µL 0.5 mm zirconia-silicate beads (Biospec, Bartlesville, OK) and subject to bead-beating according to the following program: cycles = 4, 30 sec of cycling time, speed = 6 m/s, and dwell time of 10 sec. The resulting homogenate was clarified by centrifugation at 14,000 × *g* for 3 min at 4°C, and loaded into the well of an MP96 processing cartridge containing 2μL of 1mg/mL carrier RNA (Thermofisher Scientific). The MagNA Pure 96 System (Roche) was used for the extraction with the MagNA Pure 96 DNA and Viral NA Large Volume Kit and the following protocol was run: Viral NA Plasma ext lys LV protocol with 100 μL elution volume. Nucleic acids were eluted in Elution Buffer, then stored at -80°C.

At both Windsor and Winnipeg labs, raw wastewater samples (35 mL) were also processed using Nanotrap Microbiome A Particles plus Enhancement Reagent (Ceres Nanosciences, Manassas, VA, USA) with nucleic acid extraction following lab-specific protocols described above.

**qRT-PCR**

For routine surveillance at the University of Windsor, an assay targeting the measles virus (MeV) nucleoprotein (N) gene was adopted (3). Each 20 μL reaction contained 5 μL of extracted RNA (diluted 1:3, to relieve RT-PCR inhibition), mixed with 10 μL of Luna Universal Probe One-Step Reaction Mix (2×, New England Biolabs [NEB], Ipswich, MA, USA), 1μL of Luna WarmStart RT Enzyme Mix (20×, NEB), and oligonucleotide primers and probes. Primers and probes were obtained from Integrated DNA Technologies (IDT, Coralville, IA, USA) and had final concentrations of 300nM and 150nM, respectively. The sequences of all oligonucleotides used are shown in Supplemental Table 1. The reverse transcription step was performed at 55 °C for 10 min, followed by polymerase activation at 95 °C for 2 min and 45 cycles of denaturation at 95 °C for 15 sec. Annealing/extension occurred at 63 °C for 30 sec. To create a 7-point standard curve, a synthetic measles RNA control (Twist Bioscience, South San Francisco, CA, USA) was serially diluted in ultra-pure water. No template controls (NTC) and blanks yielded no amplification. The limit of detection (LOD) for this assay was 2 copies $\mu L^{-1}$ of template or 10 copies per reaction, as determined through examination of 20 replicate standard curves. Reaction inhibition was assessed using the VetMAX XENO Internal Positive Control RNA kit (Applied Biosystems Corp., Waltham, MA, USA). Inhibition was observed in wastewater samples and as such, RNA was diluted 1:3 for all reactions. Each sample was run in technical quadruplicate using a MA6000 thermal cycler (Aumintec, Richmond Hill, ON, Canada).

The concentration of Pepper Mild Mottled Virus (PMMoV) in wastewater samples was also measured with qRT-PCR. PMMoV is a pathogenic plant virus that is broadly used as a human fecal indicator (4–6). Each 20 µL reaction well contained 2.5 µL of sample,10 µL of Luna Universal Probe One-Step Reaction Mix (2×, NEB), 1 µL Luna WarmStart® RT Enzyme Mix (20×, NEB), 3.5 µL of PCR grade water and 3 µL of combined forward primer, reverse primer, and probe each at a final concentration of 200 nM. Primer and probe sequences targeting the N gene were as previously reported (7). Reverse transcription was carried out at 55 °C for 10 min, followed by enzyme activation at 95 °C for 1 min, 40 cycles of denaturation and annealing/extension at 95 °C for 10 sec and 55 °C for 30 sec respectively. A 7-point standard curve created via dilution of a custom gBlocks (IDT) was used for quantification. The PMMoV assay had an LOD of 5 copies $\mu L^{-1}$ of template RNA, calculated by evaluation of 20 replicate 7-point standard curves, corresponding to a greater than 95% probability of detection. Negative controls yielded no amplification for any assay. Reactions were run in triplicate using a MA6000 qPCR thermocycler (Aumintec).

**Digital PCR for vaccine genotype differentiation**

To measure MeV wild-type and vaccine genotypes in wastewater, a multiplex assay developed for clinical detection was employed at the NML (8). This triplex assay targets the L gene for pan-MeV detection, and the H gene for wild-type and vaccine differentiation using two locked nucleic acid (LNA) probes for respective targets. To boost sensitivity and minimize the effects of wastewater inhibition, the Pabbaraju test was optimized for use on the Absolute Q™ Digital PCR (dPCR) System (ThermoFisher Scientific) by the NML team. Briefly, the Absolute Q™ dPCR platform was used to perform one-step RT-dPCR reactions using the 1-step RT-dPCR Master Mix (4×) (ThermoFisher Scientific, A55146). For each pre-reaction (9), 2µL of template was added to 10µL of mastermix, with a 12µL total volume. In the Absolute Q™ MAP16 Plate (Thermofisher Scientific, A52865), 9µL of the pre-reaction was added to respective sample and control wells, topped with 15µL of Absolute Q™ Isolation Buffer (Thermofisher Scientific, A52730). Cycling conditions were performed as follows: 55 °C for 10 min of reverse transcription, 96 °C for 10 min of pre-heating, followed by 40× cycling of 96 °C denaturation for 5 sec and 60 °C annealing for 15 sec. The quantification formula below was used to determine the final concentration:

$$Copies/mL = C_{instrument\ (cp/uL)} \times \frac{Volume_{prereaction\ (\mu L)}}{Volume_{template\ (\mu L)}} \times \ Dilution\ Factor\ \times \frac{Volume_{elution\ (\mu L)}}{Volume_{wastewater\ (mL)}}$$

Primer and probe sequences for the triplex assay can be found in Supplemental Table 1.

**Nanopore sequencing**

To validate the identity of the amplicon obtained from N-gene of MeV, RT-qPCR products obtained from MeV-positive wastewater samples were sequenced. Firstly, the PCR products were cleaned by adding 1 volume of NEBNext® Sample Purification Beads (NEB). DNA was eluted in 15 µL Nuclease-free water and quantified using Denovix DS-11 Spectrophotometer.

Secondly, the end-prep reaction was performed employing 250-300 ng of DNA mixed with 1.75 µL UltraII End Prep Buffer, 0.75 µL UltraII End Prep Enzyme and water to a final volume of 15 µL. The reaction was incubated at 20 °C for 10 min and 65 °C for 10 min, holding at 4 °C. Barcoding of samples was carried out combining 3 µL of end-prepped sample, 2.5 µL Native Barcode (ONT, native barcoding kit 24 V14, SQK-NBD114.24), 10 µL Blunt/TA Ligase Master Mix and 4.5 µL nuclease-free water. Ligation reaction was incubated at 22 °C for 20 min and 65 °C for 10 min, followed by a hold on ice for at least 1 min. Cleaning of the ligated sample was performed by adding 0.4 volume NEBNext® Sample Purification Beads and eluting with 12 µL of nuclease-free water. The Oxford Nanopore sequencing adaptor ligation was performed employing 200 ng of barcoded DNA in 30 µL, mixed with 5 µL Adapter Mix II, 10 µL 5X NEBNext Quick Ligation Reaction Buffer (NEB) and 5 µL Quick T4 DNA Ligase (NEB). The incubation was carried out at 25 °C for 30 min. Sample was cleaned by adding 1 volume of NEBNext® Sample Purification Beads, eluted in 12 µL of Elution Buffer and quantified. 20 ng of the library was loaded onto a SpotOn Flow Cell (R10.4.1 flow cell). Data was collected along 18 hours of sequencing with MinION™.

The *FastQ* files containing the sequenced reads were analyzed using *Epi2me* desktop application (Oxford Nanopore Technologies) to determine the presence of the expected N-gene amplicon. A preestablished workflow called *Amplicon* was employed to analyze the sequencing data. A N-450 sequence of Measles virus from strain MvS/Banjarmasin.INO/42.15/S26 (Accession number, KT964119.1) was employed as reference. After initial filtering (min_read_length: 50; max_read_length: 200; min_read_quality: 8) and adapter trimming, *minimap2* was used to align the reads to the reference. Additionally, several filtered sequences were randomly selected and manually uploaded to Basic Local Alignment Search Tool (BLAST) to confirm the identity of the N gene portion amplified by qRT-PCR.

**Sanger sequencing**

Sanger sequencing was performed as previously described (10). Briefly, MeV-positive wastewater samples underwent nested RT-PCR targeting the N450 region of the nucleoprotein gene, which was selected based on WHO guidelines as the minimum sequence required for genotype determination. Amplicons were gel purified, quantified using a Nanodrop, and sequenced at the Ottawa Hospital's Stem Core facility. Genotypes were assigned through sequence alignment using BLAST and phylogenetic analysis.

**Estimation of viral shedding**

Viral shedding rates for MeV were estimated using passive samplers deployed at WRH following a previously described approach (11) with slight modifications. Since monitoring employed passive samplers (modified Moore swabs (12)), the concentration of MeV in hospital effluent could not be measured directly. Instead, MeV concentration in the colloidal/solid fractions of hospital effluent in gene copies $L^{-1}$ (gc/L) was calculated using pellet mass and a mean suspended solids value for hospital effluent (13). Passive samplers may asymmetrically capture viruses from each fraction of effluent (14). We assumed that the passive sampling device captured only solids/colloids and did not effectively concentrate/collect the liquid fraction. Thus, the concentration of MeV in the particulate associated phase was calculated as follows:

$$MeV\ Concentration\ (Solids\ \&\ Colloids) = Copies\ per\ mass\ \left({}^{gc}/_{mg}\right) \times Suspended\ solids\ \left({}^{mg}/_{L}\right)$$

We then estimated the absolute concentration of MeV in hospital effluent using the results of fractionation experiments:

$$MeV\ concentration\ (Total) = MeV\ Concentration\ (Colloid\ \&\ Solids) \times \frac{Total\ MeV\ in\ Wastewater}{MeV\ in\ Colloids + MeV\ in\ Solids}$$

The estimated total concentration was then used to calculate a shedding rate:

$$Shedding\ rate = \frac{MeV\ concentration\ (Total\ )\ \left({}^{gc}/_{L}\right) \times Effluent\ Flow\ \left({}^{L}/_{day}\right)}{Persons\ Hospitalized \times Temporal\ Overlap}$$

Flow rates were determined using hospital water use. Admission and discharge times for MeV hospitalizations were used to determine the number of hospitalized patients. The temporal overlap value was calculated using the admission and discharge times as well as passive sampler deployment times. It represents the daily overlap between potential shedding time and sampling time and corrects for the fact that hospital admission/discharge times did not always align with passive sampler deployment/collection times. Shedding rates were reported in log10 gene copies per day per person. The median estimated shedding rate, average weekly flow and average weekly MeV concentration were used to create an estimated weekly case count for LPCC sewershed.

$$Cases\ = \frac{Mean\ LPCC\ MeV\ Concentration\ \left({}^{gc}/_{L}\right) \times Mean\ Daily\ Flow\ (\frac{L}{day}) \times \frac{7\ days}{week}}{Median\ Shedding\ Rate\ (\frac{\frac{gc}{day}}{person}) \times \frac{7\ days}{week}}$$

**Partitioning of endogenous measles virus in wastewater**

To determine the partitioning of endogenous MeV in wastewater, a 4 L post-grit wastewater sample collected from the LPCC was processed into three fractions: solids, colloids, and supernatant. Each fraction was analyzed using three biological replicates (n = 9). The approach used overlapping processing steps ensuring that for each fraction separation step, all resulting phases were tested.

Samples were first settled at 4 °C for two hours. The supernatant was carefully decanted and retained, avoiding disturbance of the settled solids. To obtain the colloids fraction, 150 mL of the post-settling supernatant from each replicate was sequentially filtered through a 1.5 µm glass fiber filter (GFF) followed by a 0.45 µm mixed cellulose ester (MCE) filter (MilliporeSigma). To recover MeV from the colloids retained on the filter surfaces, 32 mL of elution buffer (0.05 M $KH_2PO_4$, 1.0 M NaCl, 0.1% Triton X-100, pH 9.2) was passed through the used filters. The resulting eluate was considered the colloids fraction and was stored at 4 °C until nucleic acid extraction, which was performed using the QIAamp Viral RNA Mini Kit (Qiagen) on a QIAcube Connect platform, according to the manufacturer's instructions.

For obtaining the solids fraction, three biological replicates of 40 mL of the settled solids were harvested and ultracentrifuged at 1,000,000 × *g* for 45 min at 4 °C. The resulting supernatant was decanted and retained. The pellet was recentrifuged at 1,000,000 × *g* for 5 min, and the remaining supernatant was collected and combined with the previous supernatant. The final pellet, representing the solids fraction, was stored at 4 °C and processed using the AllPrep PowerViral DNA/RNA Kit (Qiagen), as previously described (15).

Finally, to obtain the supernatant fraction, three biological replicates of 37 mL of the supernatant collected during the ultracentrifugation separation step were concentrated through a 50 kDa Amicon Ultra-15 centrifugal filter (MilliporeSigma, Burlington, MA) at 4,000 × *g* for 10 min, followed by two additional spins of 20 min each to process the full volume. The concentrate retained in the centrifugal chamber was considered the supernatant fraction and was stored at 4 °C until RNA was extracted using the QIAamp Viral RNA Mini Kit (Qiagen) on the QIAcube Connect platform.

To estimate the partitioning of MeV RNA across the three fractions, the total viral signal was first calculated by summing the MeV concentration in each fraction, multiplied by the corresponding mass or volume of that fraction. Specifically, the total signal was calculated as:

$$Total\ Signal = [MeV(solids)] \times Mass\ (settled\ solids) + [MeV\ (colloids)] \times Volume\ (liquid) + [MeV(supernatant)] \times Volume\ (liquid)$$

The relative contribution of each fraction to the total signal was expressed as a percentage of this sum. Importantly, the partitioning calculations were performed using three different assumptions, each providing unique insights. First, calculations were performed on a whole-sample basis, which took into account that the wastewater sample was composed of 99.96% liquid and 0.04% solids by mass. This approach reflects theoretical distribution in wastewater but can be misleading given the dominance of the liquid phase. Second, calculations were conducted using an equal-mass basis, where 1 gram of solids was compared to 1 gram of liquid to examine which matrix contains a higher concentration of viral RNA independent of volume or composition. Finally, calculations were done using an operational volume basis, which reflects the actual processing volumes commonly used in laboratory workflows—specifically, 150 mL for the colloid fraction, 37 mL for the supernatant fraction, and 0.25 g for the solids fraction. This approach informs practical decisions for method optimization by indicating which fraction and associated concentration method would likely yield the highest detectable MeV signal under standard enrichment protocols.

**Statistical methods**

Statistical analyses were performed using R (version 4.5.1) (16). MeV gene concentrations were recorded in gene copies (gc)/mL for each sample collected. Block wise averaging was used to combine sample concentrations, producing a mean value for each of the epidemiological weeks included in the study. This allowed for direct comparison with clinical data as cases were reported weekly. A log1p transformation was applied to case counts as well as raw and normalized wastewater values but a normal distribution was not achieved and thus, we relied on non-parametric methods. To understand how well WS tracks MeV cases at the community level, Spearman's correlation was carried out between the weekly number of confirmed measles cases in Leamington and the mean weekly wastewater signal at LPCC. Spearman's rho (ρ) was calculated using base R, but the presence of equivalent values in the data necessitated the reporting of asymptotic p-values. The uncertainty of rho was estimated in the *boot* package using a non-parametric bootstrap with 1000 replications (17). Spearman's rho was recalculated for each replication, and the distribution of the estimates was used to create a 95% confidence interval. To determine if wastewater surveillance for measles virus is a leading indicator, time-stepped cross correlation was carried out in R using the ccf boot function from the *funtimes* package (18). TLCC checks the correlation between two time series at different temporal lags or shifts. If the time series are most highly correlated at a lag of 0, then they are temporally aligned, and one does not predict the other. If the correlation is strongest at a value other than 0 then one time series can be considered leading while the other is considered lagging (19).

In addition to the cross-correlation analysis, a generalized linear model (GLM) was fitted to the data to determine which factors contributed to variability in the MeV signal in wastewater. Continuous predictors were standardized via mean-centering and scaled to unit variance prior to modelling fitting. PMMoV normalized MeV RNA concentration was the response variable while covariates included: flow, pH, number of vaccinations administered (weekly), confirmed clinical cases and time (epidemiological week). Wastewater temperature was omitted as a covariate since variance inflation factor (VIF) and a correlation matrix of the fixed effects indicated that time (epidemiological week) and weekly mean wastewater temperature were collinear. The GLM was fitted to the data using the *glmmTMB* package in R (20). Since the data were continuous and right skewed a gamma distribution with a log link was used. This initial model included all predictors and maximum likelihood estimation was performed with 10,000 optimizer iterations to aid convergence. Model fit was determined using simulated residual diagnostics in the *DHARMa* package in R(21). Uniformity of residuals was tested using Kolmogorov-Smirnov tests and dispersion was evaluated with standardized residual spread relative to simulations. The frequency of outliers, quantile deviations, and the presence of temporal autocorrelation of residuals were also tested using simulated residuals. Formal tests of simulated residuals were supported by a visual examination of quantile-quantile plots, histograms and ACF plots of scaled residuals.

To investigate a potential relationship between the total volume of wastewater deposited by septic haulers and MeV wastewater concentrations, a simplified gamma GLM with a log link was fitted to a shorter data set. Normalized MeV RNA concentration was the response variable while covariates were confirmed clinical cases and total volume of sewage deposited

Finally, to further explore relationships between MeV concentrations and the volume of waste delivered by septic haulers, correlations were run between MeV and normalized MeV concentrations and the septic waste volumes on a daily basis. These data were not included in the above models as case data and vaccination administrations were only available at epi-week resolution.

### Supplementary Figures

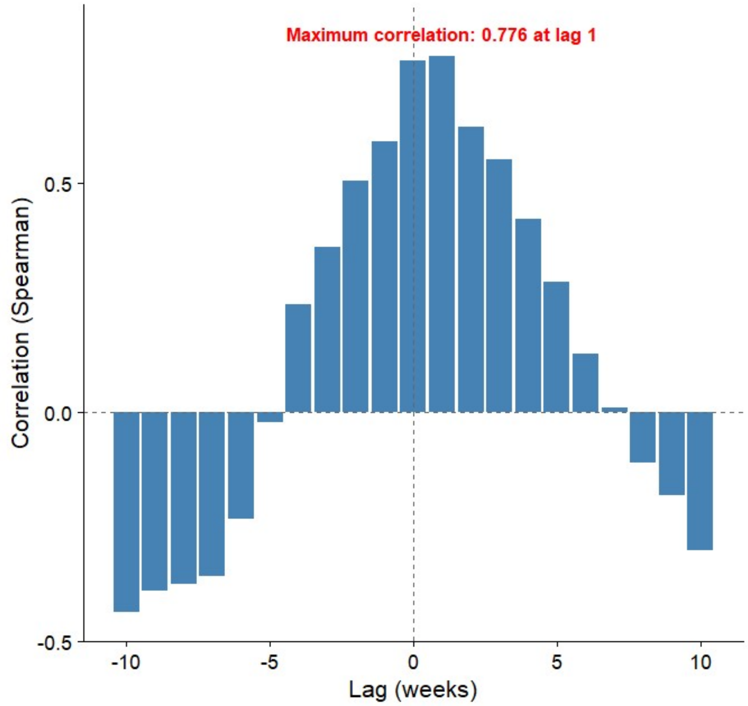


*Supplementary Figure 1. Time lagged cross correlation of weekly clinical cases of measles reported in Leamington, Ontario, Canada (2025) and a weekly average of Pepper Mild Mottle Virus normalized MeV RNA concentration in wastewater influent samples during the outbreak period (February-July 2025). Maximum correlation is found when wastewater signal lags cases by 1 week.*

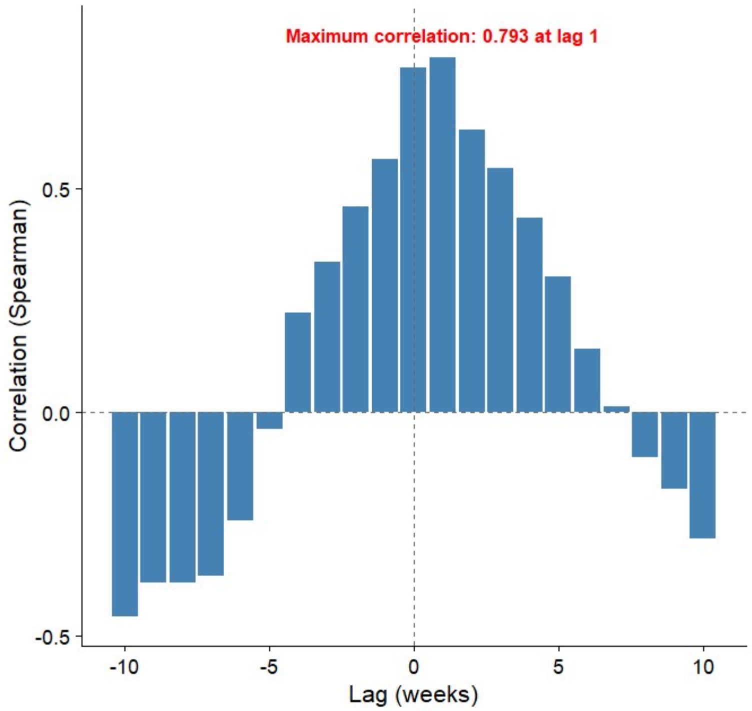


*Supplementary Figure 2. Time lagged cross correlation of weekly clinical cases of measles reported in Leamington, Ontario, Canada (2025) and a weekly average of MeV RNA concentration in wastewater influent samples during the outbreak period (February-July 2025). Maximum correlation is found when wastewater signal lags cases by 1 week.*

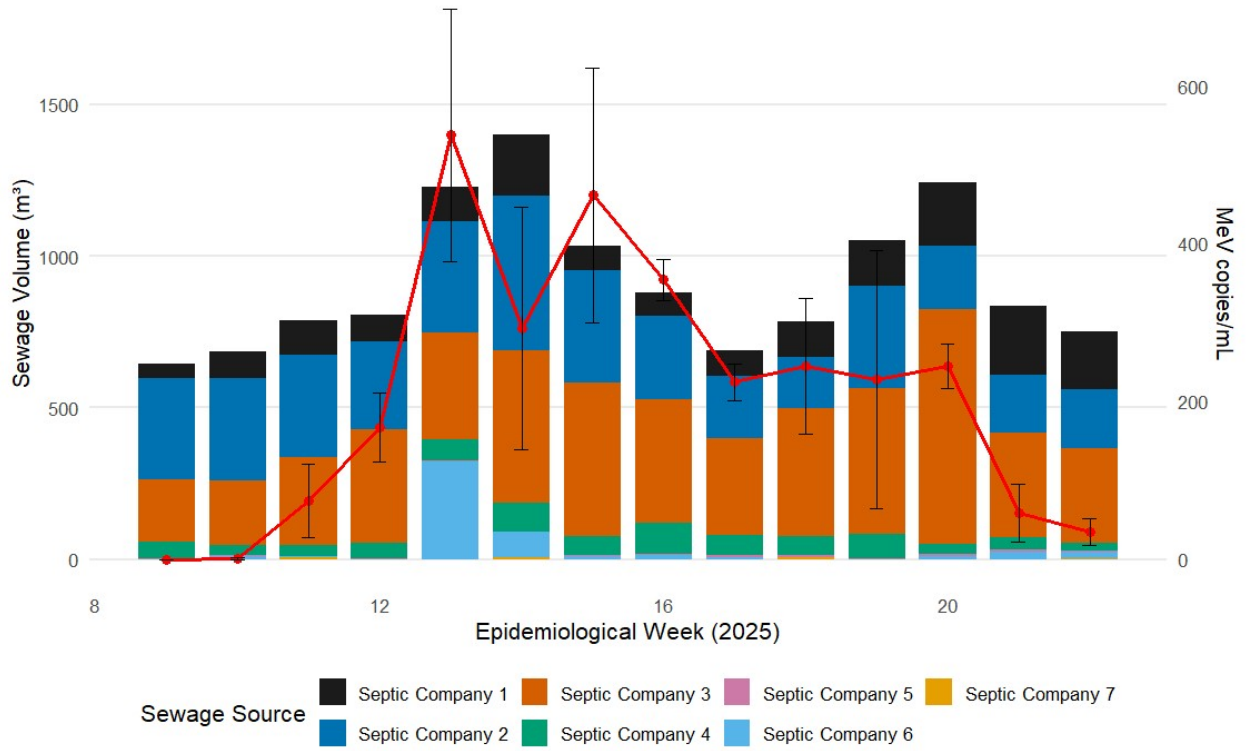


*Supplementary Figure 3. Weekly septic sewage volumes deposited by individual private septic hauling companies at the Leamington Pollution Control Centre from February to May of 2025. Stacked bars represent the sum of all sewage deposited. Superimposed on sewage volume is the mean weekly concentration of MeV RNA in gene copies per millilitre.*

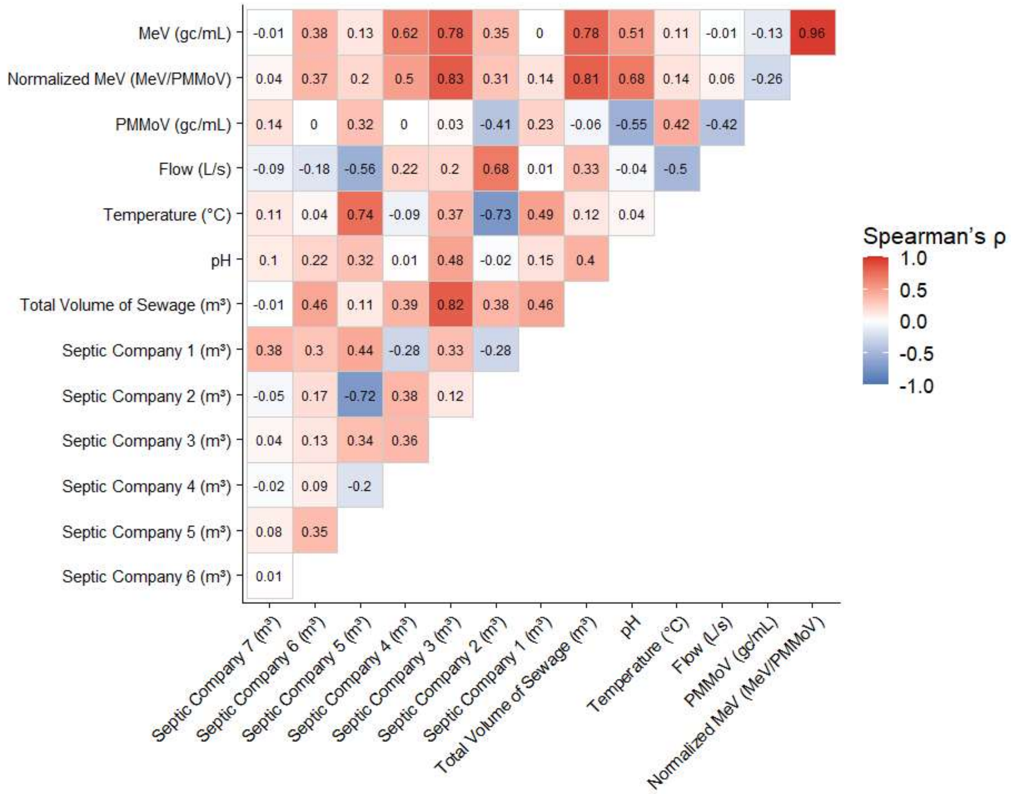


*Supplementary Figure 4. Heat map of Spearman's rho for correlations between mean weekly PMMoV normalized and raw MeV concentrations, septic sewage volumes from seven private companies, the total septic sewage volume, and physicochemical parameters of wastewater from February to May of 2025.*

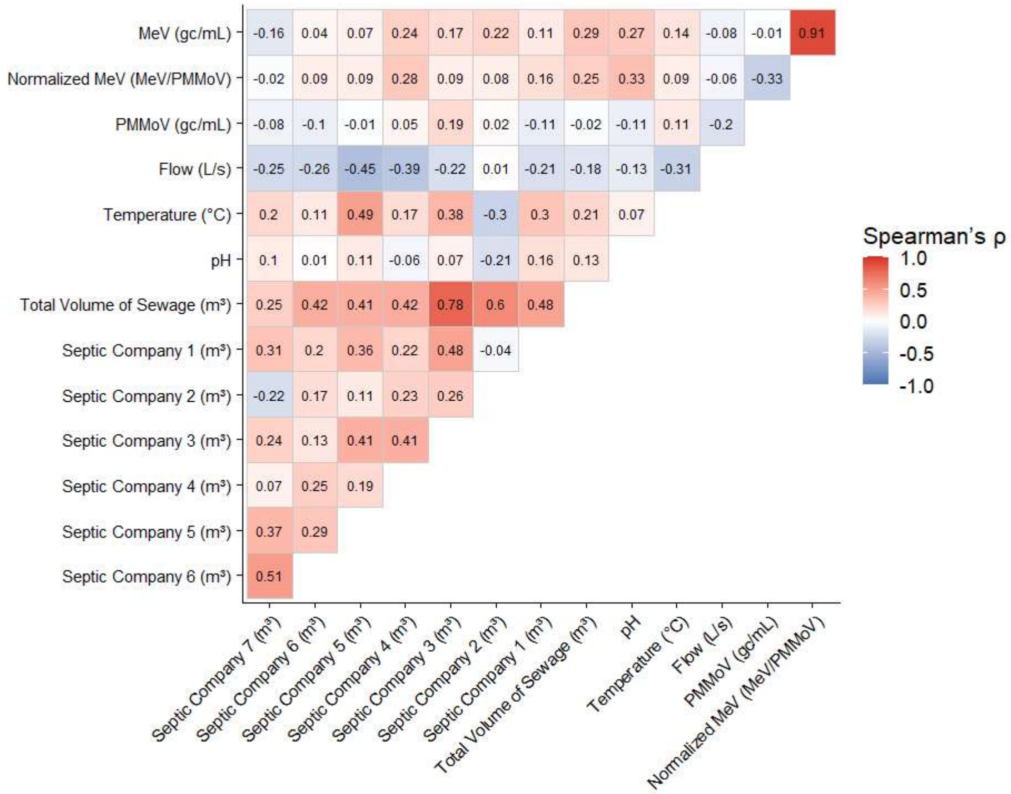


*Supplementary Figure 5. Heat map of Spearman's rho for correlations between daily PMMoV normalized and raw MeV concentrations, septic sewage volumes from seven private companies, the total septic sewage volume, and physicochemical parameters of wastewater from February to May 2025. Correlations include only data collected when septic sewage was deposited whilst composite sampling was being carried out at the wastewater treatment facility.*

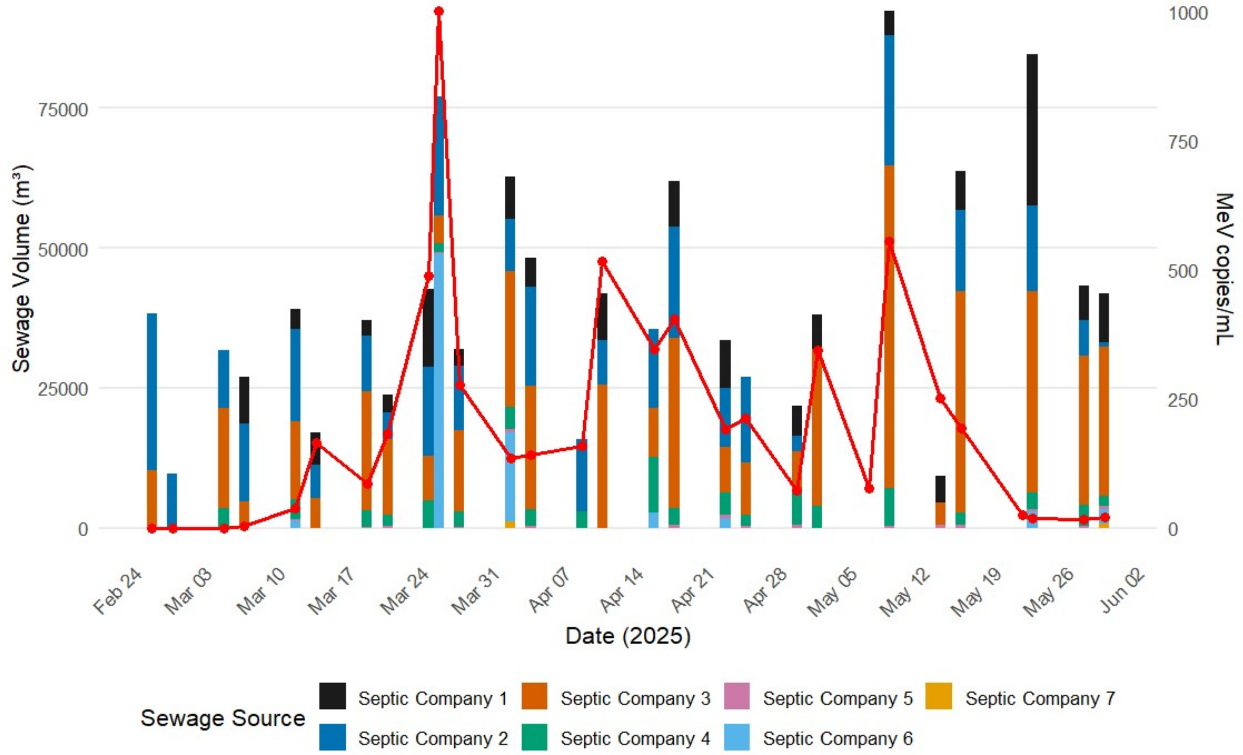


*Supplementary Figure 6. Daily septic sewage volumes deposited by individual private septic hauling companies at the Leamington Pollution Control Centre from February to May of 2025. Stacked bars represent the sum of all sewage deposited. Superimposed on sewage volume is the mean weekly concentration of MeV RNA in gene copies per millilitre. Only data collected when septic sewage was deposited whilst composite sampling was being carried out at the wastewater treatment facility is displayed.*

### Supplementary Tables

**Supplemental Table 1.** Primers and probes sequences for RT-qPCR.
[1]targets vaccine genotype

| Gene target | Primer/ probe | Sequence | Amplicon Size (kb) | Referen ce |
|---|---|---|---|---|
| N | Forward | TGGCATCTGAACTCGGTATCAC | 75 | (3) |
| | Reverse | TGTCCTCAGTAGTATGCATTGCAA | | |
| | Probe | FAM-CCGAGGATG/ZEN/CAAGGCTTGTTTCAGA | | |
| H | Forward | GAGATCCATAAAAGCCTCAGYACC | | (22) |
| | Reverse | GCCCACTTCATCMCCGATG | | |
| | Probe | Cyan500-CTAACTCAATCGAGCATCAGGTCA AGGA-BBQ | | |
| L | Forward | GCATCGAGAGAGGTTATGACCG | | (8) |
| | Reverse | TTGTGAGGAGGGGTATGACTACATC | | |
| | Probe | FAM-CTTGGCTTCACAATCA-NFQ | | |
| H1 | Forward | TGAGGACACCTCAGAGATTCACTG | | (8)<br>(8) |
| | Reverse | CCAAGTGAGATCTCTGAAGTCGTACTC | | |
| | Probe (vac) | Cy5-TGAAAT+T+ A +A-TAO-T+CT+CT+GAC-IABkFQ | | |
| | Probe (WT) | HEX-TG+AAAT+T+ C +AT+CT+CTG-IABkFQ | | |

**Supplemental Table 2.** Model diagnostic tests and for the gamma GLM fit to the full data set

| Category | Variable/Test | Estimate or Statistic | SE/Value | p-value |
|---|---|---|---|---|
| | Outcome | Normalized MeV | | |
| | Observations (N) | 23 | | |
| | Residual df | 16 | | |
| Fixed Effects | Intercept | -8.28 | 2.08 | <0.001 |
| | Weekly cases | 2.05 | 0.77 | 0.008 |
| | Flow | 0.62 | 0.41 | 0.13 |
| | pH_s | 0.43 | 0.32 | 0.17 |
| | Epi-week | 0.12 | 0.10 | 0.24 |
| | Vaccines Administered | -0.65 | 0.45 | 0.15 |
| Model Fit | Log-likelihood | 112.2 | | |
| | AIC | -210.3 | | |
| | BIC | -202.4 | | |
| Dispersion | Gamma dispersion ($\sigma^2$) | 1.23 | | |
| Residual Diagnostics (DHARMa) | Uniformity (KS test) | D = 0.13 | | 0.84 |
| | Dispersion test | 0.45 | | 0.73 |
| | Outlier test | 0 observed | Expected = 0.008 | > 0.99 |
| | Quantile deviation test | | | 0.17 |
| Temporal Dependence | Durbin–Watson statistic | 1.23 | | 0.052 |

Fixed-effect estimates are reported on the log scale with standard errors. The model was fit using a Gamma distribution with a log link. Residual diagnostics were conducted using DHARMa with 250 simulations. No evidence of non-uniformity, over- or under-dispersion, outliers, or quantile misspecification was detected. Temporal autocorrelation was marginal ($p = 0.052$) but did not exceed conventional significance thresholds.

**Supplemental Table 3.** Model diagnostic tests and for the gamma GLM fit to the truncated data set

| Category | Variable / Test | Estimate or Statistic | SE / Value | p-value |
|---|---|---|---|---|
| | Outcome | Normalized MeV | | |
| | Observations (N) | 14 | | |
| | Residual df | 10 | | |
| Fixed Effects | Intercept | -5.53 | 0.30 | <0.001 |
| | Weekly cases | 0.79 | 0.31 | 0.010 |
| | Total sewage volume | 0.90 | 0.32 | 0.004 |
| Model Fit | Log-likelihood | 57.8 | | |
| | AIC | -107.6 | | |
| | BIC | -105.0 | | |
| Dispersion | Gamma dispersion ($\sigma^2$) | 0.88 | | |
| Residual Diagnostics (DHARMa) | Uniformity (KS test) | D = 0.15 | | 0.88 |
| | Dispersion test | 0.38 | | 0.61 |
| | Outlier test | 0 observed | Expected = 0.00008 | > 0.99 |
| | Quantile deviation test | | | 0.011 |
| Temporal Dependence | Durbin–Watson statistic | 1.34 | | 0.19 |

All continuous predictors were standardized prior to modeling. Fixed-effect estimates are reported on the log scale with standard errors. Model diagnostics were assessed using DHARMa with 250 simulations. Residual diagnostics indicated adequate uniformity, dispersion, and absence of outliers. A statistically significant quantile deviation was detected; however, no evidence of temporal autocorrelation was observed.
